# Methylation-defined tumour subgroups guide recognition of genetic predisposition in Wilms tumour

**DOI:** 10.64898/2026.10.09.26364958

**Authors:** Franziska Oberhammer, Sophie E. van Peer, Nina B. Buter, Freerk van Dijk, Ronald R. de Krijger, Jette J. Bakhuizen, Reno S. Bladergroen, Martine van Grotel, Marjolijn C. Jongmans, Marry M. van den Heuvel-Eibrink, Lennart A. Kester, Roland P. Kuiper

## Abstract

Up to one-third of Wilms tumours (WTs) arise in the context of genetic or epigenetic predisposition, but identifying predisposed patients remains challenging because germline variants can be missed due to variable clinical presentation, mosaicism, and cryptic variants. We tested whether tumour-derived molecular patterns can reveal underlying predisposition and thereby complement conventional diagnostic approaches.

DNA methylation profiling of 138 WTs identified five molecular clusters that captured biological and predisposition-associated heterogeneity, including enrichment for Beckwith-Wiedemann Spectrum tumours, and a distinct *WT1*-driven subgroup. Cluster-guided reanalysis uncovered previously unrecognised *WT1* pathogenic variants, including a newly identified germline variant that led to surveillance enrolment. A random forest classifier predicted *WT1*-driven tumours with ∼90% accuracy across independent validation cohorts.

Tumour methylation profiling identifies biologically and clinically relevant WT subgroups and reliably detects a *WT1*-driven tumour methylation signature. This approach can guide targeted variant reanalysis, and uncover clinically relevant germline variants missed by standard diagnostic workflows.

## Introduction

Genetic predisposition plays a role in oncogenesis in approximately 7-12% of children with cancer^1–4^, though this percentage varies considerably across cancer types. In Wilms tumour (WT) patients, (epi)genetic predisposition has recently been described reaching substantially higher prevalence, ranging from ∼15% in earlier studies^5–7^, to up to 33% in more recent studies^8–10^. The most frequently observed predisposition syndromes in WT are Beckwith-Wiedemann Spectrum (BWSp) and *WT1*-associated predisposition, the latter encompassing conditions such as Denys-Drash syndrome and WAGR syndrome. Identifying (epi)genetic predisposition remains challenging. This is illustrated by the fact that in many patients with a high suspicion of an underlying predisposition (e.g. bilateral, multifocal, and/or familial WT) the underlying (epi)genetic aetiology remains unknown, even after intensive sequencing efforts. Contributing factors are mosaic aberrations that may be undetectable in blood^11^, and the absence of clinically recognizable features in up to 12% of genetically predisposed patients^8^. A recent study revealed that, even with current diagnostic approaches, up to 35% of patients with (epi)genetic predisposition are missed^12^. Early identification of genetically predisposed patients is important, as this enables treatment modifications, surveillance for second tumours, and counselling of family members. These considerations underscore the need for novel detection strategies.

WT is thought to arise from genetic alterations in critical nephrogenesis genes, leading to arrest during embryonic kidney development, with differentiation and/or migration being blocked^13^. These (epi)genetic events can affect one of approximately 40 WT driver genes and may arise either somatically during kidney organogenesis (spanning the differentiation of renal progenitor cells to their migration to the final anatomical position) or already be present in the germline^12^. According to Knudson’s two-hit theory, such a germline event in a tumour suppressor gene is often followed by inactivation of the remaining allele through somatic (epi)genetic events to initiate tumorigenesis^14^.

Recent studies have demonstrated that gene expression and methylation profiling can reveal subgroups with distinct molecular signatures within WT cohorts. These signatures frequently reflect dysregulation at different embryonic developmental stages and are associated with specific alterations in one or several WT driver genes^12,15–17^. We therefore hypothesised that tumour-derived molecular signatures could capture not only developmental states but also underlying (epi)genetic predisposition that may remain undetected by conventional diagnostic approaches.

In this study, we demonstrate that methylation-based profiling of Wilms tumour samples delineates distinct subgroups, reflecting underlying (epi)genetic deregulation. One subgroup is strongly enriched for *WT1* alterations, and a random forest classifier trained on methylation signatures detects this subgroup with ∼90% accuracy and high specificity. Cluster-guided reanalysis of the tumour samples uncovers previously missed pathogenic variants in five patients, including one clinically actionable germline finding. Together, these results demonstrate that tumour-based methylation profiling can complement standard germline diagnostics and improve the detection of cancer predisposition in Wilms tumour patients.

## Results

### Cohort characteristics

A total of 152 patients met the inclusion criteria and were subjected to methylation profiling. The methylation data from 138 tumours passed quality control, and the corresponding patients were included in the analyses. Median age at Wilms tumour diagnosis was 37.5 months (range 0-244 months). Biological sex was female in 75 and male in 63 patients. Median follow-up was 72 months (range 24-124 months). Full details of the cohort are available in Table **1** and Supplementary Data **1**.

**Table 1:** Baseline characteristics.

| Characteristic | Patients (n= 138) |
| --- | --- |
| **Age (months)**<br>Median<br>Range | 37.5<br>0-244 |
| **Sex**<br>Female<br>Male | 75<br>63 |
| **Laterality**<br>Unilateral<br>Bilateral | 121<br>17 |
| **Histology type<sup>b</sup>**<br>Mixed<br>Blastemal<br>Stromal<br>Regressive<br>Epithelial<br>Diffuse anaplastic<br>Non-anaplastic<sup>c</sup><br>Necrotic | 43<br>27<br>27<br>26<br>9<br>3<br>2<br>1 |
| **Preoperative chemotherapy<sup>d</sup>**<br>Yes<br>No | 129<br>9 |
| **Genetic predisposition**<br>*WT1*-related<br>BWSp<br>*DIS3L2*<br>*REST*<br>*NYNRIN*<br>Biallelic *BRCA2*<br>No<br>No consultation | 8<sup>e</sup><br>28<sup>f</sup><br>4<sup>f</sup><br>1<br>1<br>2<br>92<br>3 |

According to <sup>(a)</sup> the highest stage or <sup>(b)</sup> highest histological risk group in case a patient has multiple tumours. Non-anaplastic <sup>(c)</sup> refers to treatment-naïve tissue samples lacking preoperative chemotherapy and showing no features of anaplasia. Preoperative chemotherapy status <sup>(d)</sup> is reported at patient level. Some tumour samples were collected before treatment (see Supplementary Data 1). *WT1*-related predisposition <sup>(e)</sup> includes one additional patient identified in this study and not by standard-of-care diagnostic testing. One patient carried both a germline *DIS3L2* pathogenic variant and a BWSp diagnosis, and is therefore counted in both groups <sup>(f)</sup>. Source data are provided as a Source Data file.

(Epi)genetic predisposition for Wilms tumour was identified in 42 patients by standard of care diagnostic approaches. Twenty-seven of these patients had also been included in a previous study^8^ (Supplementary Data **1**). Among the predisposed patients, 28 were diagnosed with BWSp based on clinical criteria, with molecular confirmation in 23 patients. Pathogenic or likely pathogenic heterozygous germline variants in *WT1* were identified in seven patients, including one with WAGR syndrome. One additional patient was newly identified with a germline *WT1* pathogenic variant in this study and will be described in detail later, bringing the total number of predisposed patients to 43. A patient with a germline *REST* variant was the only patient with a positive family history for Wilms tumour.

### DNA methylation profiling identifies five distinct Wilms tumour clusters

After quality control filtering, 138 tumour samples were analysed using unsupervised consensus clustering of the 5,000 most variable CpG sites, revealing five distinct methylation-based clusters organized into two overarching groups (comprising two and three clusters, respectively) (Figure **1a**). The methylation-derived clusters demonstrated strong robustness across multiple analytical approaches. Principal component analysis of matched RNA-seq data (n=116) showed separation of samples according to DNAm cluster assignments (Figure **1b**). Consensus clustering of RNA-seq data further supported the distinct molecular profiles identified by the DNAm clustering, given their significant concordance (adjusted rand index (ARI) = 0.38, 95% CI: 0.273–0.54; permutation p < 0.001; normalised mutual information (NMI) = 0.41, 95% CI: 0.359–0.551; permutation p < 0.001; Supplementary Figure **S1a**), indicating that the methylation-defined clusters represent true biological entities reflected at the transcriptomic level. Four of the five DNAm clusters showed clear transcriptional counterparts, with clusters 1b, 2a, and 2b showing the strongest RNA-seq correspondence (71%, 72% and 88%, respectively), while clusters 1a and 2c showed moderate alignment (40% and 33%, respectively) (Figure **1c**), suggesting biological heterogeneity within this subgroup. Furthermore, DNAm clusters 1a and 1b were not only closely related by methylation but also showed partial overlap in transcriptomic clustering (Figure **1b,c**).

**Figure 1.**
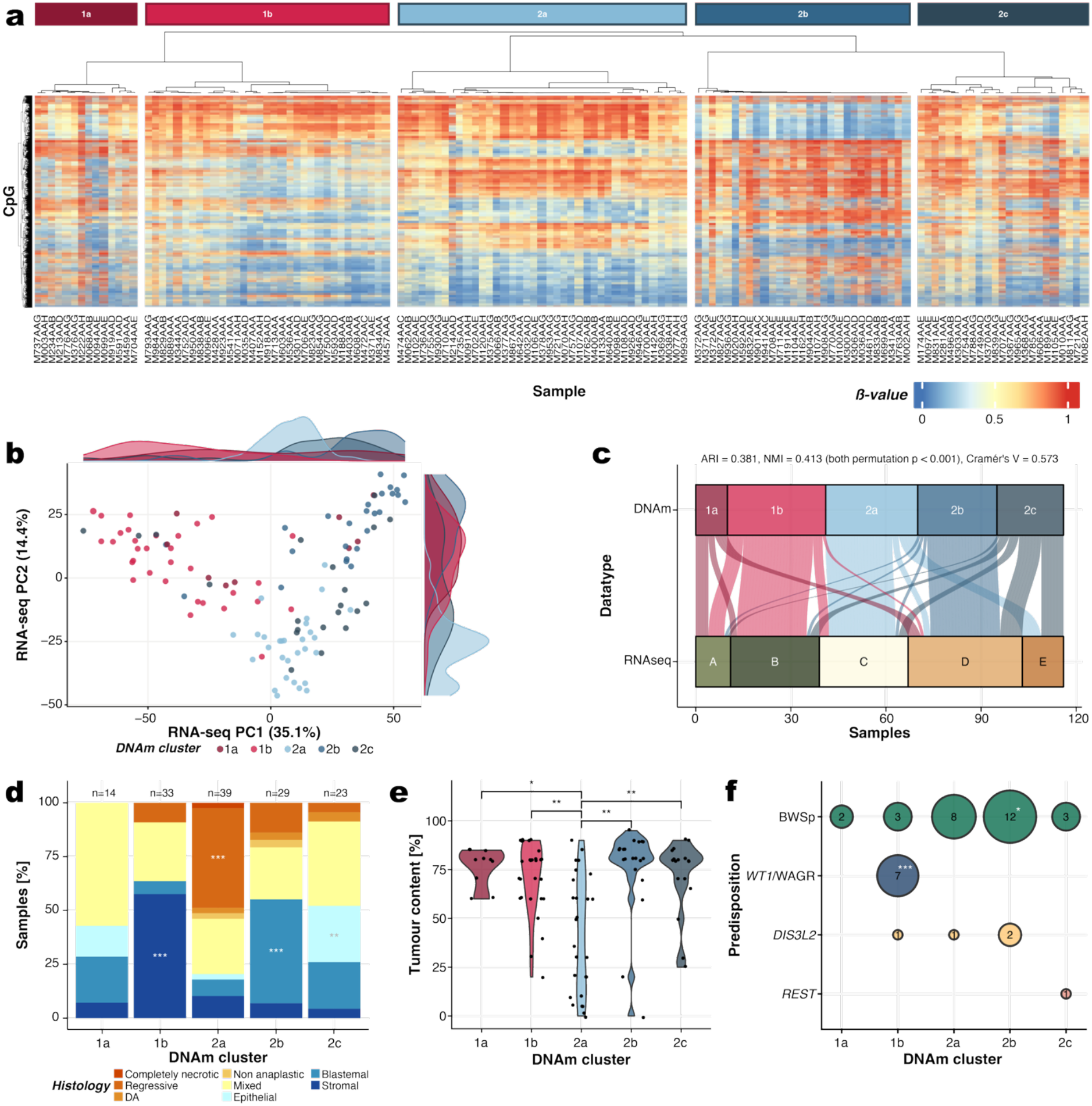
Molecular characterization of the five DNA methylation–defined Wilms tumour cluster. **a)** Heatmap showing five molecular clusters in a cohort of 138 Wilms tumours. Tumour samples (columns) and CpGs (rows) were clustered using an unsupervised consensus approach using the 5,000 most variable CpGs, as determined by standard deviation. **b)** Principal component analysis of RNA-seq data using 500 most variable genes (after correction for sex and batch), with samples coloured by DNA methylation-based cluster assignment (1a = dark red (n=14), 1b = light red (n=33), 2a = light blue (n=39), 2b = medium blue (n=29), 2c = dark blue (n=23)). Marginal density plots show cluster distribution along PC1 (top) and PC2 (left) axes. **c)** Cross-platform comparison of DNA methylation- and RNA-seq-based cluster assignments showing concordance between epigenetic and transcriptomic classifications. Clustering agreement was quantified using Adjusted Rand Index (ARI), Normalized Mutual Information (NMI) and Cramer’s V to assess association strength between categorical assignments, with statistical significance assessed by permutation testing (n=1,000 iterations). **d)** Relative frequency of histological patterns and **e)** tumour content across DNAm clusters. Statistical significance was assessed using chi-squared test (histology) and pairwise Wilcoxon rank-sum test (tumour content), adjusted for multiple testing using Bonferroni. **f)** Distribution across DNAm clusters of tumours from patients with Beckwith-Wiedemann Spectrum (BWSp), pathogenic germline variants in *WT1*, *DIS3L2*, and *REST*. Statistical significance was assessed using a chi-squared test comparing predisposition-associated patients with unaffected patients, followed by post hoc pairwise 2×2 chi-squared tests comparing each DNAm subgroup with all other subgroups. Reported p-values are Bonferroni-adjusted. *: p < 0.05, **: p < 0.01, ***: p < 0.001. Source data are provided as a Source Data file.

### DNAm clusters represent biologically distinct tumour subtypes

To understand the biological characteristics captured by the methylation profiles, we investigated the distribution of histological subtypes, CNV patterns, tumour cell content and cancer predisposition syndromes across clusters. Cluster 1b was significantly enriched for stromal histology (19/33, 57.58%; p < 0.001; standardized residuals shown in Supplementary Figure **S1b**), while cluster 2b was dominated by blastemal tumours (14/29, 48.28%; p < 0.001), and cluster 2c showed enrichment of epithelial histology (6/23, 26.09%; p = 0.001) (Figure **1d**). Diffuse anaplastic tumours were rare (one case each in clusters 2a, 2b, and 2c), while mixed histology tumours were distributed across all subgroups. Copy number variation patterns marked differences in genomic stability across clusters (Supplementary Figure **S1c-g**). Cluster 1a and 2b exhibited the highest genomic instability, characterized by frequent 1q gains (28–43% and 27-41%, respectively) and 16q losses (21–43% and 24-45%, respectively), both established poor-prognosis markers.

Cluster 2a was predominantly driven by low tumour cell content and regressive histology, displaying a significantly higher proportion of regressive samples (18/39, 46.2%; p < 0.001; Figure **1d**) and significantly lower tumour cell content (median 42.5%) compared to all other clusters (each with a median of 80%; p < 0.001; Figure **1e**). This admixture of normal kidney tissue may explain several other distinctive features of this cluster, including high chromosomal stability (occasional chromosome 12 gains in 20% of samples, Supplementary Figure **S1c**). The low tumour cell content may limit the detection of CNVs, contributing to a more normal-like methylation profile. Thus, cluster 2a does not represent a distinct biological tumour subtype but rather a sampling or tissue composition effect.

Among the WT predisposition groups represented in our cohort, *WT1*-associated predisposition and Beckwith Wiedemann Spectrum (BWSp) were the only groups with sufficient numbers to assess their distribution across DNAm clusters (Figure 1**f**). All patients with germline *WT1* pathogenic variants (7/7, 100%; p < 0.001) as well as the WAGR syndrome patient clustered in subgroup 1b (detailed variant information in Supplementary Data **2**). Tumours from patients with BWSp were found in multiple subgroups, but were most prevalent in subgroup 2b.

In summary, unsupervised methylation-based clustering reveals four biologically robust Wilms tumour subgroups (1a, 1b, 2b, 2c) characterized by distinct CNV patterns and histological features, while cluster 2a represents a mixed entity with reduced tumour content. In addition, the observed associations with WT predisposition suggest that DNAm subgroup assignment may capture features related to the underlying predisposing alterations. We therefore examined the associations with BWSp and *WT1* in more detail.

### Beckwith–Wiedemann Spectrum and tumour-specific imprinting defects

Tumours from patients with BWSp were most frequent in DNAm subgroup 2b (12/28 BWSp-associated tumours, p = 0.04). Furthermore, of the 29 tumours in this subgroup, 12 were BWSp-associated (41.4%), compared with 16 of 109 tumours across the other subgroups (14.7% p = 0.016, Figure **2a**).

**Figure 2.**
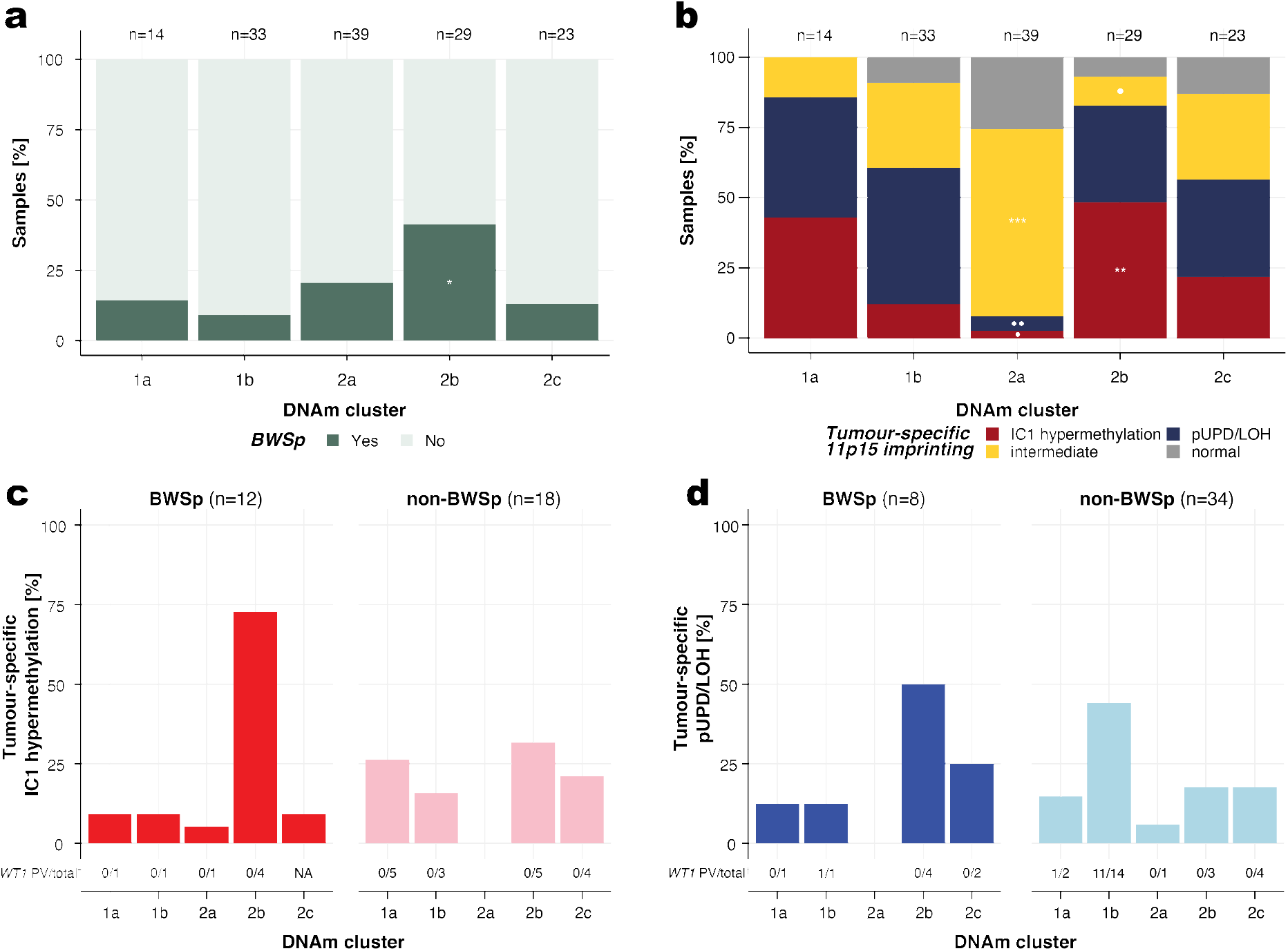
Beckwith–Wiedemann Spectrum (BWSp) and tumour-specific imprinting patterns. **a)** BWSp frequency across DNAm clusters (Fisher’s exact test with post hoc pairwise comparisons). **b)** IC1/IC2 imprinting status across DNAm clusters: hypermethylated (red), paternal UPD or LOH (blue), intermediate (yellow) and normal (grey). Sample numbers per cluster are shown above; asterisks (*) indicate significant enrichment of that specific imprinting status compared to other clusters, dots (•) indicate depletion. */•: *p*< 0.05, **/••: *p*< 0.01, ***/•••: *p*< 0.001; **c-d)** BWSp vs non-BWSp distribution across DNAm clusters, stratified by imprinting status (hypermethylation and pUPD/LOH, respectively). Intermediate and normally imprinted samples were removed from this analysis. Numbers below each bar indicate the number of samples with a pathogenic variant (PV) in *WT1* over the total number of samples with available sequencing data (†). *WT1* data were unavailable for 6 samples in the hypermethylated group and 10 samples in the pUPD/LOH group. Source data are provided as a Source Data file.

In our cohort, BWSp patients presented with two main categories of underlying 11p15 alterations, namely either hypermethylation of the IC1 locus alone, or combined IC1 hypermethylation with IC2 hypomethylation due to paternal uniparental disomy (pUPD) in this region. These patterns were also reflected in the tumours (Supplementary Figure **S2a**). Importantly, similar 11p15 methylation patterns can also result from tumour-acquired events in sporadic WT, including somatic pUPD and loss of heterozygosity (LOH)^53^. As these two events produce comparable methylation patterns, they could not be distinguished based on methylation alone. Nevertheless, IC1 hypermethylation and pUPD/LOH methylation patterns were clearly distinguishable in the tumours (30 and 42 cases, respectively, Supplementary Figure **S2b**). We therefore analysed the distribution of these two tumour methylation patterns across DNAm subgroups separately. A subset of tumours (n=48) showed intermediate imprinting patterns that could not be attributed to either of these groups or to normal imprinting. Their enrichment in cluster 2a likely reflects higher normal cell content in this subgroup rather than true imprinting defects (Figure **2b**, Supplementary Figure **S2c-d**). Overall, IC1-hypermethylated tumours were enriched in cluster 2b (14/29, 48.3%; p = 0.002, Figure **2b**, Supplementary Figure **S2c**). This pattern was largely driven by BWSp-associated tumours, which clustered predominantly in 2b, whereas, non-BWSp tumours were more broadly distributed across subgroups (range: 5%-32%) (Figure **2c**, Supplementary Figure **S2e**). The distribution of tumours with 11p15 pUPD/LOH was more heterogenous. Most pUPD/LOH tumours in subgroup 1b harboured biallelic *WT1* alterations, suggesting that the 11p event in these tumours reflects WT1-driven tumour evolution rather than BWSp. BWSp-associated pUPD/LOH tumours, however, were most frequently assigned to subgroup 2b, similar to those with IC1 hypermethylation (Figure **2d**, Supplementary Figure **S2e**).

Together these findings suggest that DNAm subgroup assignment may help identify tumours associated with underlying BWSp, particularly in the context of IC1 hypermethylation, although it does not allow definite distinction from somatically acquired 11p15 methylation defects.

### *WT1* variant detection through clustering-guided analysis

The enrichment of germline *WT1* PVs in subgroup 1b led us to re-evaluate tumours in this subgroup without a previously detected *WT1* variant, through thorough inspection of *WT1* expression, splice junctions, copy-number profiles, and variant calls. By doing so, we newly identified *WT1* alterations in five patients that were missed by standard diagnostics: four patients in whom no *WT1* variant had previously been detected, and one patient in whom a second hit was identified. The alterations comprised a somatic exon 1 deletion followed by CN-LOH (patient M035AAD), duplication of parts of *WT1* exon 7 (M706AAE), two chromothripsis events resulting in partial or complete loss of *WT1* (patients M407AAA and M593AAD), and one intronic variant predicted by SpliceAI to affect splicing (patient M371AAE) (Figure **3a**, Supplementary Data **2**). This latter patient was of particular interest because of stromal-predominant histology of the tumour, intralobar nephrogenic rests and young age at diagnosis, features strongly suggestive of *WT1*-associated predisposition. At diagnosis, the variant (GRCh38 NC_000011.10(NM_024426.6):c.1016+89G>A) was not called by WES pipelines. After clustering-guided reanalysis of WGS and RNA-seq data, the variant was found to be heterozygous in peripheral blood and normal kidney, but was homozygous in the tumour due to CN-LOH of 11p (Figure **3b**). SpliceAI predicted disruption of the canonical donor splice site and creation of a novel splice site, consistent with the intron retention observed in tumour transcriptomic data. A minigene assay subsequently confirmed that the variant causes aberrant splicing, resulting in intron retention (Figure **3c**). This germline variant was found to be *de novo* in this patient and, following its identification, the patient was placed under surveillance consistent with *WT1*-associated cancer predisposition management guidelines.

**Figure 3.**
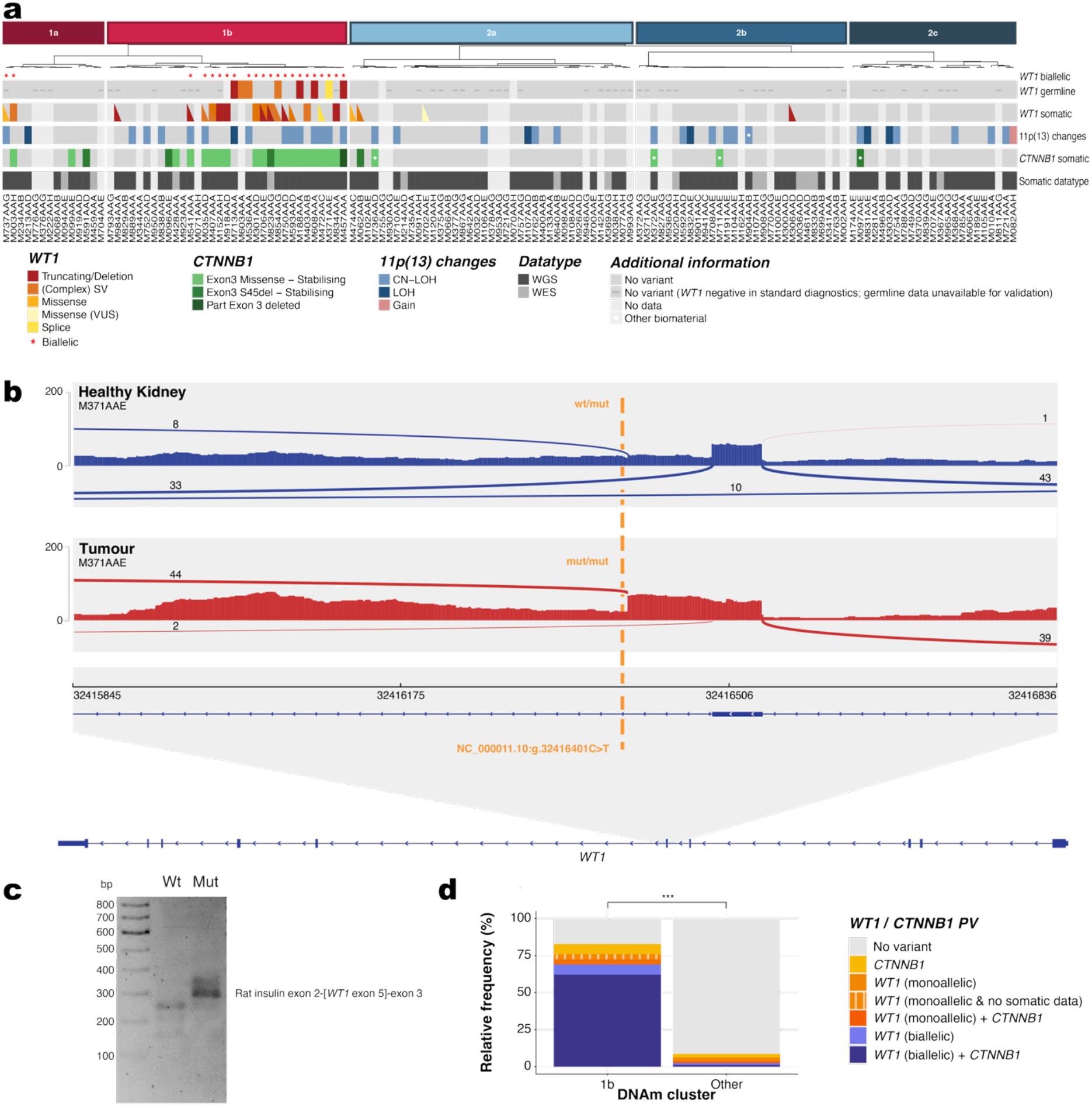
Characterization of *WT1* and *CTNNB1* alterations in Wilms tumour cohort. **a**) Overview of *WT1* and *CTNNB1* alterations in the Wilms tumour cohort. Samples are ordered by global methylation cluster (Figure 1). *WT1* alterations: truncating variants or deletions (red), (complex) structural variants (orange), missense variants (light orange), splice variants (yellow) and missense variants of uncertain significance (light yellow). (CN-)LOH affecting (parts of) chromosome 11p is indicated in blue (light and dark blue, respectively); copy-number gains are indicated in light red. An asterisk (*) denotes cases with biallelic *WT1* alteration. *CTNNB1* alterations: stabilizing exon 3 missense mutations and S45 deletions, and partial exon 3 deletions (VUS) are shown in progressively darker greens. A white dot denotes a mutation detected in a different biomaterial than that used for methylation profiling. Dark grey indicates absence of the respective alteration; light grey, no data. Datatype: Somatic mutation detection was performed by WGS (dark grey) or WES (light grey); the germline *WT1* PV in patient M371AAE and somatic PVs in patients M035AAD, M407AAA, M593AAD, as well as M706AAE (second hit) were not identified by standard-of-care diagnostic testing. Details about all alterations can be found in Supplementary Data **2**. **b)** RNA-seq coverage plot for a tumour of a patient with a newly detected germline intronic *WT1* variant (GRCh38 NC_000011.10(NM_024426.6):c.1016+89G>A; yellow dotted line=variant position). The intronic variant in intron 5 generates a cryptic splice site 4 bp upstream and weakens the canonical splice site 89 bp upstream, resulting in intron retention after exon 5. Normal kidney shows low-level usage of the cryptic site; tumour shows predominant cryptic-site usage caused by CN-LOH. **c**) Molecular validation of aberrant *WT1* splicing in healthy kidney (blue) and tumour tissue of patient M371AAE. Minigene splicing assay using a rat insulin exon 2-[*WT1* exon 5]-exon 3 reporter construct, resolved by agarose gel electrophoresis; the variant-carrying sample shows a longer band (∼300 bp) consistent with aberrant splicing and intron retention. **d**) Relative frequency of *WT1* and *CTNNB1* mutations in cluster 1b versus all other clusters. Samples were excluded if no somatic data were available: 1b = 26 samples, other = 74 samples, ***:*p*< 0.001, determined by chi-squared test. Source data are provided as a Source Data file.

### Somatic and predisposition-associated *WT1* alterations converge in subgroup 1b

In addition to the eight tumours from patients with germline *WT1* PVs, 20 tumours harboured somatic *WT1* variants, of which 14 also clustered in subgroup 1b. Thirteen of these 14 tumours showed biallelic *WT1* inactivation (Figure **3a** and Supplementary Data **2**). Whereas the second hit in predisposition-associated tumours was consistently acquired through (CN-)LOH of 11p13, frequently extending to 11p15.5, second hits in tumours with somatic *WT1* variants were more heterogeneous and included both (CN-)LOH and secondary mutations.

Of the six *WT1* variant-harbouring tumours outside subgroup 1b, two showed biallelic inactivation and were assigned to subgroup 1a, the DNAm subgroup closely related to 1b. The remaining four tumours were assigned to subgroups 2a and 2b. The *WT1* variant in one of these tumours was classified as VUS. Furthermore, none had a detectable second hit and were therefore not considered *WT1*-driven. This illustrates how DNAm subgroup assignment may aid in contextualising the pathogenic relevance of *WT1* variants.

*CTNNB1,* the most frequently co-mutated gene in *WT1*-altered tumours^54^, showed a similarly strong association with *WT1* status and subgroup assignment in our cohort. Among the 21 *WT1*-altered tumours in subgroup 1b with available somatic sequencing data, 18 (85.7%) harboured a co-occurring somatic *CTNNB1* variant, compared to 2/6 (33.3%) outside subgroup 1b. Overall, subgroup 1b showed significant enrichment for *CTNNB1* variants (20/28, 71%) relative to other subgroups (4/81, 4.94%; p < 0.001). The majority (23/24) were classical gain-of-function mutations located in exon 3, which encodes critical phosphorylation sites that regulate β-catenin degradation and thus Wnt pathway activation^55^; the remaining variant was a partial deletion spanning parts of intron 2 and exon 3.

Taken together, subgroup 1b is characterised by enrichment of germline and somatic *WT1* alterations, observed in 75.9% of subgroup 1b tumours versus 6.17% in all other subgroups combined (Figure **3d**, p < 0.001), establishing *WT1*-associated predisposition and somatic alterations as defining molecular features of this subgroup.

### Development of a DNA methylation–based multi-class classifier for Wilms tumour subgroups

To enable classification of samples into the DNAm-defined subgroups, we developed a supervised random forest classifier trained on the 10,000 most variable CpG sites across DNAm clusters (Figure **4a**), adapting the MnM minority classification framework for DNAm data^34^. Internal performance was evaluated by 10-fold cross-validation, yielding high overall performance (balanced accuracy 88%, specificity 96%, sensitivity, precision, and F1 all >79.7%, Figure **4b–c**, Supplementary Figure **S3a**). Performance was strongest for the *WT1*-enriched subgroup 1b (balanced accuracy = 95%), while subgroup 1a showed reduced stability and lower classification performance, (balanced accuracy = 78%), consistent with a less homogeneous epigenetic profile that was not resolved by adjusting cluster granularity. Furthermore, overall performance improved when low-confidence predictions (<73% probability) were excluded (balanced accuracy = 91%, sensitivity = 84%, specificity = 97%, F1 = 86%, confusion matrix can be found in Supplementary Figure **S3b**).

**Figure 4.**
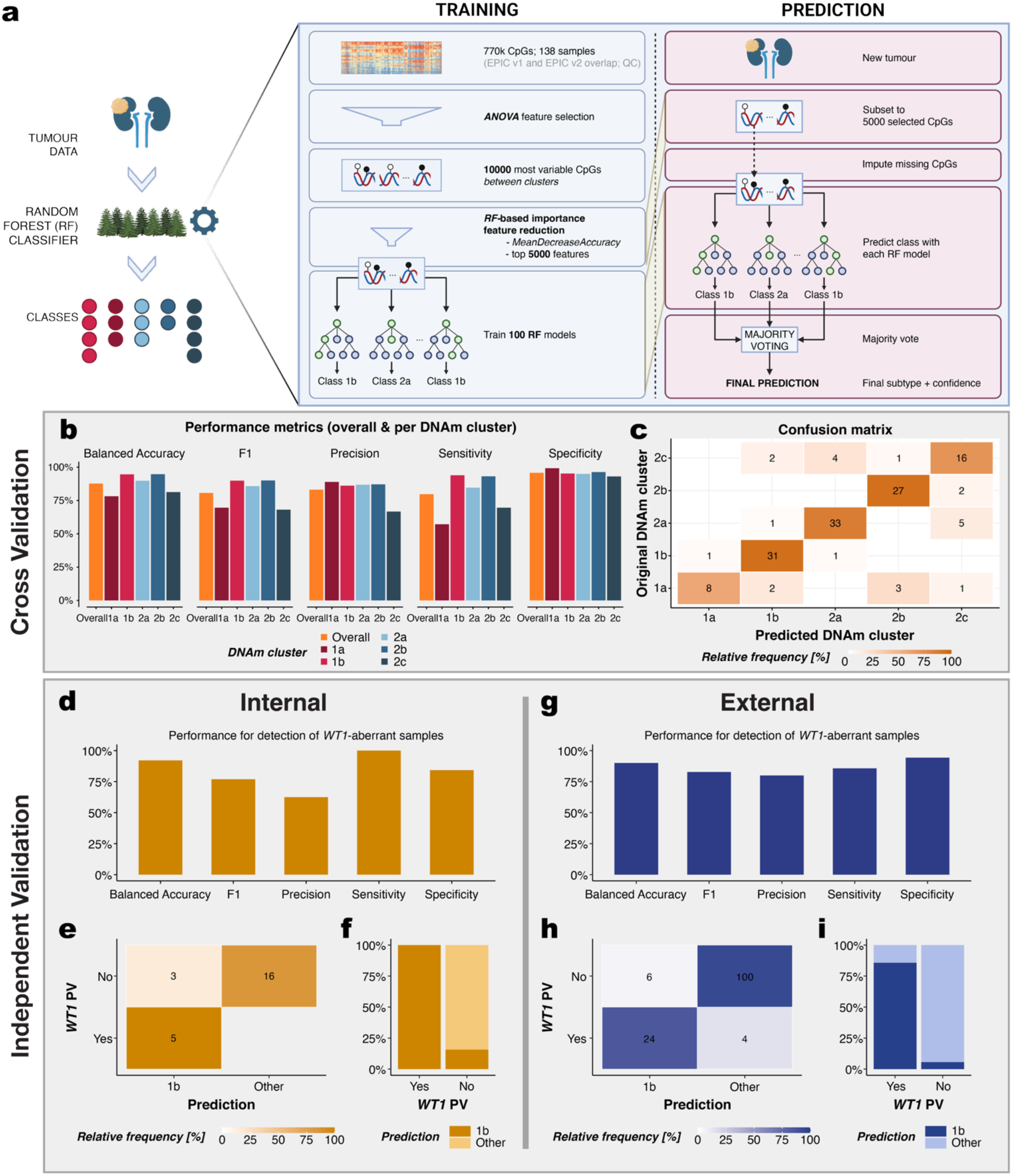
Classifier training, validation, and performance. **a)** Framework of classifier training and prediction (created with BioRender). **b–c)** Classifier performance in cross-validation: **(b)** overall and per-class performance metrics, with corresponding confusion matrix **(c)**. **d–i)** Performance in independent validation cohorts: overall metrics for internal **(d)** and external **(g)** validation on *WT1* PV detection, with corresponding confusion matrices **(e)** and **(h)**, respectively. Stacked bar plots showing the percentage of samples belonging to cluster 1b (dark) or other classes (light) among samples with and without *WT1* alterations of the internal (**f**) and external (**i**) validation cohorts. Source data are provided as a Source Data file.

### Validation on independent cohorts confirms classifier generalizability

The final classifier model was applied to two independent validation cohorts. Ground-truth DNAm cluster labels were unavailable for independent cohorts. Classifier performance was therefore evaluated based on its ability to identify *WT1*-associated tumours, given their strong enrichment in subgroup 1b. For the internal validation cohort, *WT1* status was comprehensively assessed through SNV, SV, and CNV analysis, including (CN-)LOH, and *WT1* expression levels relative to the whole cohort (detailed variant information in Supplementary Data **3**). One tumour (M953AAF) lacking an identifiable PV in *WT1* by genomic analysis was nonetheless found to harbour CN-LOH of chromosome 11 and showed near-absent *WT1* expression, a pattern comparable to tumours with confirmed *WT1* loss where residual expression reflects normal kidney tissue only. Given this evidence, this tumour was classified as harbouring a *WT1* PV. For the external cohort, *WT1* status labels as provided by the original study were used directly.

Within this internal validation cohort (n=24), balanced accuracy reached 92%, with 100% sensitivity and all known *WT1*-altered tumours correctly classified as subgroup 1b (5/5, Figure **4d–f**). In the external validation cohort (n=134), the binary classification of *WT1*-associated versus non-*WT1*-associated patients showed high performance, with balanced accuracy of 90% and specificity of 94% (Figure **4g**). Patients with *WT1* alterations were predominantly assigned to subgroup 1b (24/28, 85.7%), and those without *WT1* alterations to non-*WT1* subgroups (100/106, 94.3%) (Figure **4h-i**). Of the 10 discordant assignments, four were false negatives (Supplementary Figure **S3c**). One patient with a germline *WT1* PV was classified into subgroup 2a, the cluster characterised by low tumour content and regressive tumour samples. Consistent with this classification, all analysed tumour samples from this patient showed low tumour content, as inferred from IC1 and IC2 imprinting status (Supplementary Figure **S3d**). Two patients were classified into subgroup 1a, the DNAm subgroup most closely related to 1b, with one showing a classification probability below the confidence threshold, indicating an uncertain assignment. The fourth patient carried both a somatic *WT1* mutation and kidney-mosaic 11p15 loss of imprinting; despite the somatic *WT1* alteration, this patient was classified into subgroup 2b, where BWSp cases predominate. No clinical data were available for the external cohort to determine whether this patient meets BWSp criteria. The six samples without a reported *WT1* variant may either lack an identifiable *WT1* alteration, as we also observed for some subgroup 1b tumours in our training cohort, or harbour an alteration that was not detected in the original study. This could not be further assessed because sequencing data were unavailable.

Together, these results demonstrate that the DNAm-based classifier robustly identifies *WT1*-associated Wilms tumours across independent cohorts, supporting its potential utility for detecting previously unrecognised *WT1*-related predisposition in clinical and research settings.

## Discussion

This retrospective study demonstrates that methylation-based clustering identifies biologically, genetically and histologically distinct Wilms tumour (WT) subgroups, including patterns associated with underlying (epi)genetic predisposition. In particular, subgroup 1b was strongly associated with *WT1* alterations, while BWSp-associated tumours were preferentially represented in subgroup 2b. Cluster-guided analysis enabled the identification of previously missed *WT1* alterations, and a DNAm-based classifier allowed reproducible subgroup assignment and robust identification of *WT1*-associated tumours across independent cohorts. Together, these findings build on previous evidence that Wilms tumour subgroups reflect distinct biology and that predisposition can shape tumour development^12,16^. We show that these molecular features can also be used to recognize underlying predisposition and guide targeted genetic re-evaluation.

Germline *WT1* PVs were exclusively detected in subgroup 1b, in line with previous studies showing that *WT1*-driven Wilms tumours follow a characteristic molecular trajectory and display distinct molecular profiles^12,56^. Frequent co-occurring somatic *CTNNB1* alterations in tumours with biallelic *WT1* inactivation further support subgroup 1b as a molecularly defined entity^55,57^. Importantly, our findings extend these observations by showing that DNAm subgroup assignment may also be used in the reverse direction: to flag tumours in which an underlying *WT1* alteration should be considered. Cluster-guided re-evaluation of subgroup 1b tumours without a previously detected *WT1* variant uncovered five additional variants missed by standard diagnostics, including chromothripsis events and a germline intronic splice-altering variant, directly leading to appropriate clinical surveillance in this patient. This suggests a potential future diagnostic workflow in which a *WT1*-associated methylation profile prompts targeted re-analysis of tumour and germline data for missed alterations, including cryptic, structural, and deep-intronic variants.

The enrichment of BWSp-associated tumours in DNAm subgroup 2b suggests that methylation-based classification captures features associated with BWSp-related tumorigenesis. However, this association was less specific than for *WT1*-associated tumours, as BWSp-associated tumours were also distributed across other subgroups. This heterogeneity could partly be explained by additional tumour characteristics, including *WT1* alterations and low tumour cell content, which may influence subgroup assignment. Importantly, tumours with late somatically acquired 11p15 methylation alterations showed a different subgroup distribution than BWSp-associated tumours. These findings suggest that combining subgroup assignment with tumour imprinting status may provide additional information when underlying BWSp is considered, although this approach cannot independently distinguish BWSp-associated from sporadic 11p15 alterations, and will require integration with clinical and constitutional testing.

To enable reproducible and prospective DNAm subgroup assignment, we developed a random forest classifier and validated it in internal and external independent cohorts. This validation confirmed that the classifier can reliably identify *WT1*-associated tumours across datasets from different centres, supporting the feasibility of applying it to samples from newly diagnosed WT patients for prospective classification. One patient with a germline *WT1* PV in the external validation cohort was misclassified into subgroup 2a due to low tumour content, highlighting the importance of adequate tumour purity for reliable classification. Formal validation for BWSp status was not feasible as neither independent cohort contained patients with confirmed BWSp, and will require dedicated validation in other cohorts in future studies. Thus, the potential value of classifier-based subgroup assignment for BWSp recognition remains to be established. More generally, the classifier provides a reproducible approach for prospectively assigning newly diagnosed tumours to the DNAm-defined subgroups and using these molecular patterns to guide further evaluation of underlying predisposition.

This study has several limitations mostly originating from its retrospective design. Non-uniform sample processing and incomplete datasets were unavoidable. Data from normal kidney tissue was available for only a subset of patients, preventing assessment of 11p15 imprinting status in certain cases and potentially leading to missed (mosaic) BWSp diagnoses. Tumour sampling may not fully capture intratumoral heterogeneity, though we addressed this by re-evaluating histology based on sampled areas. In addition, a considerable number of samples clustered in subgroup 2a, which was characterized by low tumour content and regressive features, underscoring the importance of obtaining material with sufficient tumour content. One potential approach to address this is to computationally mix DNA methylation signals from defined tumour clusters with normal kidney profiles and retrain a secondary classifier. In exploratory analyses, this strategy enabled reclassification of some 2a samples. Further optimisation and validation will be needed before this approach can be incorporated into the classifier, because the underlying tumour subgroup of low-tumour-content samples cannot be independently established.

Sample sizes were limited by the rarity of individual predisposition conditions, restricting the present analysis to *WT1* and BWSp. Although these represent the most common predisposition types in WT^8,9,12^, extending the approach to additional WT predisposition genes will be important. Looking ahead, incorporating more tumours from patients with pathogenic variants in other predisposition genes such as *TRIM28* and *DIS3L2* would broaden the scope of the classifier; *TRIM28*-associated tumours represent a distinct epithelial subtype with genome-wide methylation deregulation^16,58,59^, that typically lack additional driver mutations^56^ and may define an additional methylation-based subgroup. Additionally, applying methylation profiling to cell-free DNA could enable earlier, less invasive, predisposition diagnosis and improved guidance of treatment and recurrence surveillance. This would be particularly relevant for patients with *WT1* PVs, in whom the risk of chronic kidney disease and kidney failure further increases the importance of nephron-sparing surgery.

In conclusion, this study establishes methylation profiling as a promising approach for recognizing underlying (epi)genetic predisposition in Wilms tumours, particularly *WT1*-associated predisposition, and suggests potential value for identifying BWSp-associated tumours. DNAm subgroup assignment can guide targeted genetic analysis towards specific WT-predisposing genes and loci, and uncover clinically relevant (germline) variants missed by standard testing. As shown by the patient in whom a germline *WT1* PV was identified upon methylation-guided tumour reanalysis, integration of somatic tumour profiling with germline assessment could improve the identification of patients with underlying predisposition and thereby support better patient care and family counselling.

## Methods

### Patient selection & ethics statement

Patients with a Wilms tumour diagnosed between January 2015 and July 2023 with informed consent for the Princess Máxima Center Biobank study (Netherlands trial register NL7744, IRB approval number MEC-2016-739), SIOP-RTSG WT 2001 study (EudraCT number 2007-004591-39, ethics approval number MEC 202.134/2001/122) and/or SIOP-RTSG-2016 UMBRELLA study (ethics approval number MEC-2018-026) were eligible for inclusion in the training cohort. Fresh frozen tumour material was required for inclusion. For internal validation, tumour material from 24 additional patients with available fresh frozen tumour material and informed consent for the Princess Máxima Center Biobank study and/or SIOP-RTSG-2016 UMBRELLA study were included. For external validation, tumour methylation data from 134 patients from the GSE292896 dataset were included. This study was conducted in accordance with Good Clinical Practice guidelines and the Declaration of Helsinki.

### Sample preparation and histological re-examination

DNA and RNA were isolated from fresh frozen tumour samples and germline DNA was isolated from whole blood leukocytes and sequenced according to the institute’s standard procedures^18^. Because Wilms tumours often display mixed histology, an expert paediatric tumour pathologist re-examined H&E-stained sections adjacent to the DNA/RNA sampling site and reassigned histologic subtype for each specimen. This sample-level reclassification ensured that molecular analyses were interpreted in the correct histological context.

### Genetic predisposition diagnostics

Genetic predisposition classification was based on comprehensive germline DNA sequencing data collected through standardized diagnostic protocols. As standard of care, all patients with Wilms tumour were referred to clinical genetics for comprehensive phenotypic and genetic assessment. This included whole exome sequencing (WES) with subsequent analysis of a targeted panel of 29 WT predisposition genes^19^. Molecular diagnostic criteria followed current consensus guidelines for variant classification^20^. In addition, Methylation-Specific Multiplex Ligation-dependent Probe Amplification (MS-MLPA) was performed in a diagnostic setting to detect 11p15 methylation defects in blood-derived DNA. For patients presenting with clinical features indicative for Beckwith-Wiedemann Spectrum (BWSp) but negative MS-MLPA results in blood, MS-MLPA results from healthy kidney cells were included when results were available^8^.

Annotation of patients with an (epi)genetic predisposition was based either on the gene harbouring a pathogenic or likely pathogenic germline variant, or on a clinically and/or molecularly confirmed syndromic diagnosis.

### Sequencing data processing and genomic characterisation

Pre-processing of whole transcriptome sequencing (RNA-seq), whole-genome sequencing (WGS) and whole-exome sequencing (WES) data was performed using the institution’s standardized pipelines, which implement GATK 4.0 best practices using a wdl and cromwell-based workflow^21^. A minimum median coverage of 30x for normal and 90x for tumour WGS data was used, while for WES data the minimum median coverage was 100x and 200x, respectively. A minimum of 30 million unique reads was used for RNA-seq data. Quality control was performed according to established institutional criteria^18^.

Somatic variant calling for tumour-normal pairs was carried out using *Mutect2* (GATK v4.2.0.0^21^) and GRCh38 reference genome according to GATK best practices. When only tumour tissue was available, variant calling was performed using GATK practices with GATK *HaplotypeCaller*. The resulting variant calls were annotated with Ensembl VEP v105^22^. Single nucleotide variants (SNVs), multi-nucleotide variants, and small insertions/deletions (InDels) that passed the ‘PASS’ filter were further filtered based on quality criteria (depth ≥ 10, alternative allele count ≥ 5 and variant allele frequency ≥ 0.1) and population frequency (GnomAD v3.0 ≤ 1%)^23^. Variants predicted to be pathogenic were retained if they had a High Impact consequence, a CADD_Phred score ≥ 15, and/or a SpliceAI score ≥ 0.5^24,25^. Afterwards the remaining variants were manually assessed using Integrative Genomics Viewer (IGV)^26^.

Structural variants (SVs) were identified using two separate approaches. When tumour and germline data were both available for a sample, an in-house developed SV pipeline was used to generate a merged call set from the output of three callers^27,28^. The call set was filtered to include only variants with a filter value of “PASS”, a tool count ≥ 2 and normal AF ≥ 0.1. If only tumour data was available, calling was performed using *MANTA* v1.6.0^29^in tumour-only mode. Only “PASS” filter SVs supported by both paired and split reads were considered, with thresholds of PR_ALT ≥ 0.1 and PR_TOT ≥ 10 or SR_ALT ≥ 0.1 and SR_TOT ≥ 10. For both approaches SVs longer than 10,000 bp and/or with at least one breakpoint within a gene were selected for further analysis. All chosen SVs were visually inspected and validated using IGV.

Copy number variation (CNV) analysis was performed from WGS, WES or, when sequencing data was unavailable, using DNAm data. The package c*onumee2* was used to infer CNV status on DNAm. Raw intensity data from Illumina EPIC array v1 and v2 were imported and pre-processed using *minfi* and harmonized to GRCh38 reference genome using the respective annotation packages (v1: *IlluminaHumanMethylationEPICanno.ilm10b5.hg38*, v2: *IlluminaHumanMethylationEPICv2anno.20a1.hg38*). Preprocessing included Illumina control probe normalization and background correction, followed by extraction of methylated and unmethylated intensities. Only probes present on both array versions were retained for cross-platform compatibility.

CNV detection used a set of 50 randomly selected control samples from GSE246337 as copy-number neutral references^30^. CNVs were inferred using the *conumee2* package. Summary CNV plots were generated for previously defined methylation-based clusters using *CNV.summaryplot*. Only alterations present in at least 20% of the samples within a cluster are reported.

### DNA methylation profiling and clustering

DNA methylation profiles were assessed using Illumina Human MethylationEPIC BeadChip arrays (v1.0 and v2.0) at the molecular pathology department of the University Medical Center Utrecht according to the manufacturer’s instructions. Data import, quality control (QC) and preprocessing were performed using the Bioconductor package *SeSAMe* with the settings QCDPB. QC metric included number of probes, signal detection, signal intensity, colour channel and dye bias. For each metric, samples were assessed relative to the cohort mean, with values within ±3 standard deviations (SD) considered valid; samples exceeding 10% of QC measures outside this range were excluded. Bisulfite conversion efficiency was evaluated using array-specific probes, and samples outside ±3 SD of cohort were removed from further analysis. Successful background subtraction was confirmed by visual inspection of the ß-value distribution by probe design, and dye bias correction was validated using Red-Green QQ plots. ß-values were calculated, and for EPIC v2.0 samples, CpGs targeted by multiple probes were collapsed using the method *minPval* in *getBetas.* Only CpGs present on both array versions were retained to ensure cross-platform compatibility. Poor quality probes were masked using *SeSAMe’*s design quality mask for v1.0; for v2.0 masking was performed manually using the *getMask* function, as automated masking was not yet supported at the time of analysis. Methylation of probes targeting SNPs was used to detect potential sample swaps, and biological sex was predicted using *inferSex;* samples flagged for either swaps or sex discordance were excluded.

To correct for unwanted technical variation, batch effects and confounding covariates (i.e. sex) were removed using *limma::removeBatchEffect.* To identify which covariates require correction, principal component analysis (PCA) was first performed using the R package *sta*t. The principal components cumulatively explaining ≥ 80 % of total variance were examined for correlation with technical and biological covariates, and those showing significant association were included in the batch correction model.

The 5,000 CpGs sites showing the highest variability across samples were selected for clustering analysis using the Bioconductor package *ConsensusClusterPlus* with *innerLinkage* and *finalLinkage* method *ward.D2*, agglomerative hierarchical clustering algorithm, *pearson* as distance method with 1,000 re-samplings and 80% sample subsampling. Cluster stability was assessed for different numbers of clusters (*k*=2-16) using the consensus matrix, the consensus cumulative distribution function plot and the delta area plot. The optimal *k* was selected based on the point where further increasing *k* no longer substantially improved cluster stability. Clustering results were visualized using the package *ComplexHeatmap*.

Annotation files for Illumina Infinium MethylationEPIC BeadChip arrays, versions 1 and 2, were obtained from the InfiniumAnnotation resource maintained by Zhou and colleagues^31^. The annotation resource was accessed in May 2024.

### Imprinting centre methylation analysis

Methylation analysis of Beckwith-Wiedemann spectrum (BWSp) associated imprinting centres (ICs) was performed using probes within IC1 (chr11:1998149-1998777, GRCh38) and IC2 (chr11:2699310-2700164, GRCh38) regions, corresponding to the CTCF binding sites previously reported by Silva *et al.*^32^. ß-values from batch-corrected methylation data were extracted for probes present on both EPIC v1 and v2 arrays, combined into a single matrix, mean-centred and scaled. Consensus clustering was performed using *ConsensusClusterPlus* with *Ward’s linkage*, *hierarchical clustering*, and *Euclidean distance*. Cluster stability was assessed as described above to identify the optimal cluster number. Results were visualized using *ComplexHeatmap* with consensus clustering dendrogram and annotation tracks.

### Statistical analyses

Associations between DNA methylation clusters and categorical variables were assessed using a two-step approach. First, a contingency table was constructed for each variable versus cluster membership. For sparse tables (≤10 cells), Fisher’s exact test was applied; for larger tables, a chi-squared test with simulated p-values (10,000 simulations) was used to account for low expected counts. Only if the overall test was significant (p ≤ 0.05) additional per-cluster analyses were performed. For chi-squared tests, standardized residuals were computed for each cluster-category combination, and Bonferroni correction was applied to identify significantly enriched or depleted cells. For Fisher exact tests, each cluster was compared against all other clusters using 2×2 tables, with p-values adjusted for multiple testing. Standardized residuals were used to visualize deviations from expected distributions, with positive values indicating enrichment and negative values indicating depletion. Combinations with adjusted p-values ≤ 0.05 were considered significant and highlighted in the visualization. Differences in tumour content percentage between DNA methylation clusters were evaluated using pairwise Wilcoxon rank-sum tests. P-values were adjusted for multiple testing using the Bonferroni method.

### Transcriptome analysis

RNA-seq count data were processed using DESeq2 and converted to a DESeqDataSet object with experimental design accounting for batch effect and biological sex. Low-expressed genes were filtered by retaining genes with ≥10 read counts in at least 10% of samples. Size factors were estimated using the median-of-ratio method to normalize for sequencing depth differences. Samples with extreme library sizes (>3 standard deviations from the mean total normalized counts) were identified and removed from downstream analysis. Variance stabilizing transformation (VST) was applied to stabilize variance across the mean expression range. For PCA, batch effects (sequencing batch and sex) were removed from VST-transformed data using *limma::removeBatchEffect*. The top 500 most variable genes were selected by row variance and PCA was performed using *prcomp*. Variance explained was calculated as the proportion of total variance per component.

Consensus clustering was performed on batch-corrected VST data using ConsensusClusterPlus with hierarchical clustering (Ward.D2 linkage), Pearson correlation distance, and 1,000 iterations with 80% item sampling. The optimal cluster number was determined using the consensus cumulative distribution function and delta area metrics. Cross-platform concordance between DNA methylation and RNA-seq clustering was assessed using the Adjusted Rand Index (ARI) and Normalized Mutual Information (NMI), with significance evaluated by permutation testing (n=1,000) and 95% bootstrap confidence intervals. Associations between clustering assignments were tested using chi-squared tests with Monte Carlo simulations for small expected frequencies.

### *WT1* and *CTNNB1* variant analysis

*WT1* and *CTNNB1* variants were obtained from processed variant calling data. All germline variants were classified according to the ACMG criteria^20^. Pathogenicity of somatic variants was classified using ClinVar or, when unavailable, Franklin. Variants of unknown significance (VUS) were distinguished from pathogenic variants (PV) and labelled accordingly. Biallelic *WT1* inactivation was determined per sample by considering any combination of germline PV, somatic PV, (copy-neutral) loss of heterozygosity, or multiple somatic variants; samples meeting these criteria were marked with asterisks in the oncoplot. To further characterise WT1 alterations at the transcript level, RNA-sequencing read coverage across the *WT1* locus was visualized using sashimi plots generated in IGV for paired healthy kidney and tumour samples from the same patient.

### Splice assay (minigene)

To assess the functional impact of a newly identified *WT1* variant (GRCh38 NC_000011.10(NM_024426.6):c.1016+89G>A) on splicing, a minigene assay was performed using the pSpliceExpress vector system^33^. This allows the splicing behaviour of the inserted sequence to be assessed in a controlled cellular context independently of endogenous regulatory elements. Both the wild-type and mutant sequences were cloned and transfected in parallel to directly compare splicing outcomes and attribute any observed aberrant splicing to the variant.

The genomic region spanning *WT1* exon 5 and flanking intronic sequences (GRCh38 chr11:32,415,977–32,416,919) was amplified from patient germline DNA using a nested PCR approach with TaKaRa Ex Premier DNA Polymerase (Clontech, RR370A). In the first PCR, gene-specific nested primers were used to amplify the region of interest (94°C 1 min; 35 cycles of 98°C 10 s, 61°C 15 s, 68°C 25 s). The product was gel-excised and purified using the QIAquick Gel Extraction Kit (Qiagen), and used as template in a second PCR with attB-tailed adapter primers under identical cycling conditions to enable Gateway cloning (primer sequences in Supplementary Data **4**).

The purified attB-flanked PCR product was cloned into pSpliceExpress by BP recombination using the Gateway BP Clonase II Enzyme Mix (Invitrogen) following the manufacturer’s instructions, transformed into Stellar chemically competent cells (Takara Bio), plated on LB-Ampicillin agar (50 µg/ml). Individual colonies were expanded overnight in LB-Ampicillin and screened by colony PCR using M13 forward and reverse primers (94°C 1 min; 35 cycles of 98°C 10 s, 60°C 15 s, 68°C 25 s, sequences provided in Supplementary Data **4**). Positive clones were confirmed by Sanger sequencing (Macrogen) following ExoSAP-IT treatment, and plasmid DNA was purified using the NucleoBond Xtra Maxi Kit (Macherey-Nagel) following the manufacturer’s instructions.

HEK293FT cells were seeded at 7×10⁵ cells per well in 6-well plates in high-glucose DMEM with GlutaMAX and transfected 24 h later with sequence-verified wild-type and mutant pSpliceExpress constructs using PEI in Opti-MEM. An empty pSpliceExpress backbone and a GFP plasmid served as controls. After 48 h, cell pellets were harvested and total RNA was isolated using the Maxwell RSC SimplyRNA Cells Kit (Promega) including on-column DNase I treatment, following the manufacturer’s instructions; RNA concentration was quantified using the QuantiFluor RNA System (Promega). cDNA was synthesised from 1 µl RNA per sample using the High-Capacity cDNA Reverse Transcription Kit (Applied Biosystems) with random primers in a 20 µl reaction, following manufacturer’s instructions. Splicing products were amplified by RT-PCR using vector-specific primers targeting the flanking rat insulin 2 exons (Rat-Ins2-exon2-F / Rat-Ins2-exon3-R; sequences in Supplementary Data **4**) with TaKaRa Ex Premier DNA Polymerase (94°C 1 min; 34 cycles of 98°C 10 s, 57°C 15 s, 68°C 25 s). DNA fragments were separated via agarose gel electrophoresis, stained with SYBR Green (Bio-Rad), and gel images were acquired using the GelDoc XR+ imaging system (Bio-Rad). To confirm the sequence identity of the amplification products, target bands were excised and purified using the QIAquick Gel Extraction Kit (QIAGEN) for subsequent Sanger sequencing.

### DNA methylation-based classification

An ensemble-based Random Forest classifier was developed using the *MnM* minority class modelling approach to classify samples into one of the five identified methylation subtypes^34^. The classifier development consisted of the following steps.

First, ANOVA-based feature selection was performed to identify the most informative CpG sites for classification. A one-way ANOVA was performed comparing M-values across the five methylation subtypes, and the top 10,000 CpG sites with the highest F-statistics were selected as candidate features for model training.

Second, one hundred Random Forest models were trained independently to reduce model-specific bias (*randomForest::randomForest*). For each model iteration, a balanced training set was generated by sampling from each methylation subtype with probability inversely proportional to the square root of class frequency, ensuring representation of all subtypes. Any underrepresented methylation subtype missing from the random sample was supplemented by adding one additional sample from that class. Samples appearing more frequently than a specified maximum threshold (determined as the median number of samples per subtype) were down sampled. Feature reduction was performed to identify the most important CpG sites using Random Forest feature importance, with the top 5,000 features (by Mean Decrease in Accuracy) from each model retained. Each Random Forest model was trained using 500 decision trees with Mean Decrease in Accuracy as the feature importance metric. Class weights were inversely proportional to class frequency to address class imbalance, and bootstrap sampling was employed.

Third, the feature importance values across all 100 models were aggregated to identify the most consistently important CpG sites. The 5,000 top-ranked features were selected based on their mean importance scores across all models. These reduced feature sets were used to train the final ensemble of 100 Random Forest models with identical parameters.

Finally, sample classification was performed via majority voting. Predictions from all 100 models were aggregated by counting votes for each class, with the subtype receiving the most votes designated as the prediction. A classification confidence score was calculated as the percentage of models voting for the majority class (range 0-100%).

### Classifier validation

The classifier’s performance was evaluated using 10-fold cross-validation on the training cohort. The training cohort was randomly divided into 10 non-overlapping folds while maintaining balanced representation of all five methylation subtypes in each fold. For each fold, models were trained on the nine remaining folds and tested on the held-out fold. This process was repeated 10 times, yielding predictions for all samples in the training cohort. Classification accuracy was calculated as the proportion of correctly classified samples. Confusion matrices were generated to assess per-subtype classification performance. For each methylation subtype, sensitivity (true positive rate), specificity (true negative rate), precision (positive predictive value), F1 score (harmonic mean of precision and recall), and balanced accuracy (mean of sensitivity and specificity) were calculated using the *caret* package. Classification confidence was furthermore evaluated by filtering predictions based on confidence thresholds. Samples with confidence scores ≥ 73% were considered high-confidence predictions, while those below this threshold were classified as low-confidence.

Independent internal validation samples, separate from the training cohort, were classified using the trained classifier. The same preprocessing steps (EPIC array processing, beta-to-M-value conversion) were applied to maintain consistency with training data. Methylation values at imprinting control regions 1 and 2 on chromosome 11 were assessed as quality metrics and indicators of imprinting status.

An external validation cohort from an independent study and institution was used to assess classifier performance on external data. Samples from the external validation cohort underwent identical preprocessing and classification procedures. Classification predictions and confidence scores were analysed to determine whether the classifier generalizes to samples processed outside the training laboratory.

### Software

| <u>Package</u> | <u>Version</u> | <u>Purpose</u> |
| --- | --- | --- |
| sesame<sup>35,36</sup>/minfi<sup>37,38</sup> | 1.23.9/1.48.0 | DNA methylation array preprocessing and analysis |
| limma<sup>39,40</sup> | 3.58.1 | Statistical modelling |
| sva<sup>41,42</sup> | 3.50.0 | Batch-effect correction |
| ConsensusClusterPlus<sup>43,44</sup> | 1.66.0 | Consensus clustering |
| MnM<sup>34</sup> | 1.0.1 | Original MnM classifier framework adapted for DNA methylation data |
| randomForest<sup>45</sup> | 4.7-1.1 | Random forest classification |
| umap<sup>46</sup> | 0.2.10.0 | Dimensionality reduction |
| ComplexHeatmap<sup>47,48</sup> | 2.18.0 | Heatmap visualization |
| ggplot2<sup>49</sup> | 3.4.4 | Data visualization |
| DESeq2<sup>50,51</sup> | 1.42.1 | RNA-seq data processing |
| conumee2<sup>52</sup> | 2.1.2 | Copy-number analysis from methylation array data |

## Data availability

The datasets generated and analysed during the current study are available upon request in the European Genome-phenome Archive (EGA) under accession number EGAS00001008535. Source data are provided with this paper.

Publicly available DNA methylation data used for CNV analysis can be accessed through the Gene Expression Omnibus (GEO) under accession number GSE246337. Additionally, DNA methylation data used to validate the random forest classifier are available through GEO under accession number GSE292896.

## Code availability

All custom code and scripts developed for this study are publicly archived on Zenodo (DOI: <u>10.5281/zenodo.22791270</u>) and maintained on GitHub. Standard bioinformatic tools and pipelines used in the analysis are cited in the Methods section. Researchers wishing to classify their own samples are welcome to contact the corresponding author.

## Artificial intelligence disclosure statement

AI tools were used to assist with improving the clarity and presentation of the text and code, including minor language editing, formatting, and stylistic refinements. The scientific content, analyses, and final decisions were made and verified by the authors.

## Funding and acknowledgements

This work was supported by Stichting Kinderen Kankervrij (Foundation KiKa), grant No. 439 and 522 to RPK, LAK, MCJJ, and MMvdHE. The funder played no role in study design, data collection, analysis and interpretation of data, or the writing of this manuscript. Furthermore, FO is supported by HE/MSCA co-fund project no 101081481. We thank Wilbert Vermeij and Alice Larsson for providing the healthy kidney data for one of the patients in this study. We also thank the Princess Máxima Center Biobank and the Princess Máxima Center Genomics Core for their support.

## Author contributions statement

**FO** contributed to Conceptualization, Methodology, Software, Validation, Formal analysis, Investigation, Data curation, Writing (original draft, review & editing), Visualization. **SEvP** contributed to Conceptualization, Methodology, Formal analysis, Investigation, Resources, Data curation, Writing (original draft, review & editing), Visualization. **NBB** contributed to Validation, Investigation, Resources, Data curation, Writing (review & editing). **FvD** contributed to Software, Formal analysis, Investigation, Data curation, Writing (review & editing). **RRdK** contributed to Resources, Writing (review & editing). **JJB** contributed to Resources, Data curation, Writing (review & editing). **RSB** contributed to Investigation, Writing (review & editing). **MvG** contributed to Resources, Writing (review & editing). **MCJJ** contributed to Conceptualization, Resources, Data curation, Writing (review & editing), Supervision, Funding acquisition. **MMvdHE** contributed to Conceptualization, Resources, Writing (review & editing), Supervision, Funding acquisition. **LAK** contributed to Conceptualization, Methodology, Writing (review & editing), Supervision, Funding acquisition. **RPK** contributed to Conceptualization, Methodology, Writing (original draft, review & editing), Visualisation, Supervision, Project administration, Funding acquisition.

